# Full-length merozoite surface protein 1 of *Plasmodium falciparum* is a major target of protective immunity following controlled human malaria infections

**DOI:** 10.1101/2022.10.12.22280947

**Authors:** Micha Rosenkranz, Irene N. Nkumama, Sara Kraker, Marie Blickling, Kennedy Mwai, Dennis Odera, James Tuju, Kristin Fürle, Roland Frank, Emily Chepsat, Melissa C. Kapulu, CHMI-SIKA study team, Faith H. A. Osier

## Abstract

The merozoite surface protein 1 (MSP1) is the most abundant protein on the surface of the invasive merozoite stages of *Plasmodium falciparum* and has long been considered a key target of protective immunity. However, previous studies focused on small C-terminal fragments and potentially missed the opportunity to identify important epitopes that are relevant for protection. We used samples from a controlled human malaria challenge (CHMI) study in semi-immune volunteers to show that levels of pre-challenge antibodies directed against the full-length MSP1 (MSP1_FL_) are significantly correlated with protection from malaria. Furthermore, we showed that anti-MSP1_FL_ antibodies induced five distinct Fc-mediated effector mechanisms: complement fixation, phagocytosis, respiratory burst, degranulation and IFNγ production, each of which was strongly associated with protection. The breadth of Fc-mediated effector functions was the strongest correlate of protection. Our findings suggest that MSP1_FL_ is an important target of functional antibodies that contribute to a protective immune response against malaria and support the development of MSP1_FL_-based vaccines.

## Introduction

Malaria remains a serious public health concern with ∼ 241 million cases and ∼ 641 000 deaths in 2020 (WHO report 2021). The need for a highly effective malaria vaccine that offers durable protection remains urgent. The World Health Organisation (WHO) and its partners set a goal to develop a malaria vaccine with >75% efficacy by 2030 (WHO, 2018). However, the currently approved vaccine, RTS,S (Mosquirix™, GSK Bio) conferred approximately 30% efficacy and protection was relatively short-lived in a phase III clinical trial (Partnership, 2015). The RTS, S related vaccine candidate R21 has recently achieved promising results in a phase II clinical trial with >77% protection for one year (Datoo et al., 2021) which was prolonged upon boosting (Datoo et al., 2022). However, more and larger studies are needed to confirm these results.

Epidemiological observations demonstrate that humans living in malaria-endemic regions naturally acquire immunity following repeated *Plasmodium falciparum* infections (Marsh and Kinyanjui, 2006). It is widely accepted that antibodies play a key role in antimalarial immunity (Cohen et al., 1961; Sabchareon et al., 1991) and emerging data highlight the importance of Fc-mediated effector functions for a protective immune response. This includes the recruitment of complement factors (Boyle et al., 2015; Reiling et al., 2019) natural killer (NK) cells (Odera et al., 2021), monocytes (Hill et al., 2013; Musasia et al., 2022; Osier et al., 2014) and neutrophils (Joos et al., 2010). We recently conducted a controlled human malaria infection study in semi-immune Kenyan adults (CHMI-SIKA) (Kapulu et al., 2022) where we showed that antibody-dependent phagocytosis of ring-stage parasites (Musasia et al., 2022) and activity of NK cells against merozoites (Odera et al., 2021) were important functional correlates of protection. Furthermore, additional analyses incorporating the full-panel of Fc-dependent and non-Fc mechanisms revealed that the breadth of effector functions targeting merozoites was most strongly associated with protection (Nkumama et al., 2022).

Two independent lines of evidence led us to investigate full-length merozoite surface protein 1 (MSP1^FL)^ as a target of important Fc-mediated mechanisms. First, it is the most abundant protein on the merozoite surface (Gilson et al., 2006) and we have recently shown that the breadth of effector functions was strongly correlated with antibody binding to whole merozoites (Nkumama et al., 2022). Second, an independent and recent vaccine trial using MSP1_FL_ showed that the vaccine was safe, highly immunogenic and induced opsonizing antibodies that fixed complement and triggered neutrophil respiratory burst (Blank et al., 2020). In other studies, antibodies targeting MSP1 have been shown to promote several mechanisms, including direct inhibition of parasite growth by blocking RBC invasion (Blackman et al., 1990; Woehlbier et al., 2010; Woehlbier et al., 2006) or inducing Fc-mediated effector functions such as complement fixation (Boyle et al., 2015), opsonic phagocytosis activity of monocytes (Kana et al., 2019) and respiratory burst of neutrophils (Jäschke et al., 2017; Joos et al., 2015). However, only Jäschke and colleagues studied functional activity (ADRB and invasion inhibition) of antibodies against MSP1_FL_, while others focussed on subunits of MSP1 such as p42 or p19. Hence, the full repertoire of effector functions against MSP1_FL_ has yet to be analysed.

While MSP1 based vaccines showed protective efficacy in animal models (Etlinger et al., 1991; Herrera et al., 1990; Perrin et al., 1984) clinical trials in humans have been disappointing overall. Importantly, all human trials focused on subunits of MSP1 such as the p42 fragment rather than the full-length molecule and therefore missed ∼80% of the protein and important epitopes that might be relevant for protection (Ogutu et al., 2009; Sheehy et al., 2012).

The evidence that antibodies targeting MSP1 are important for naturally acquired immunity (NAI) has been conflicting. This is in part due to differences in the subunit, allelic form and quality of the protein that was assessed (al-Yaman et al., 1996; Dodoo et al., 1999; Egan et al., 1996; Nebie et al., 2008; Osier et al., 2008; Richards et al., 2013). More importantly, most sero-epidemiological studies focused on small subunits of the full-length MSP1 molecule from either the conserved C-terminal (MSP1^19^ or MSP1^42^) (Okech et al., 2004; Wilson et al., 2011) or the polymorphic N-terminal domain (MSP1 Block 2) (Cavanagh et al., 2004; Cavanagh et al., 1998). As with MSP1-based vaccines, the opportunity to identify important epitopes for protection may have thus been missed. Using MSP1_FL_ we now show that volunteers who were protected from sporozoite challenge had high levels of anti-MSP1_FL_ antibodies and induced MSP1_FL_-specific Fc-mediated effector function in five distinct assays involving complement, neutrophils, natural killer cells and phagocytes. Our findings suggest that the previous narrow focus on the C-terminal subunits of MSP1 may have been misplaced and elevate the full-length molecule as an important target of protective immunity against malaria.

## Results

### Anti-MSP1_FL_ antibody levels were higher in non-treated versus treated CHMI-SIKA volunteers

To test the potential relevance of MSP1_FL_ in protection from malaria, we assessed MSP1_FL_-specific IgG, IgM and IgG subclass antibodies in plasma samples from CHMI-SIKA volunteers (n=142) that were collected one day before sporozoite challenge (C-1). Interestingly, we found that the seroprevalence of IgG was comparable in treated and non-treated volunteers (95% versus 100%, respectively), while IgM was predominant in non-treated volunteers (69% versus 20%, respectively, **Fig. 1A**).

**Figure 1:**
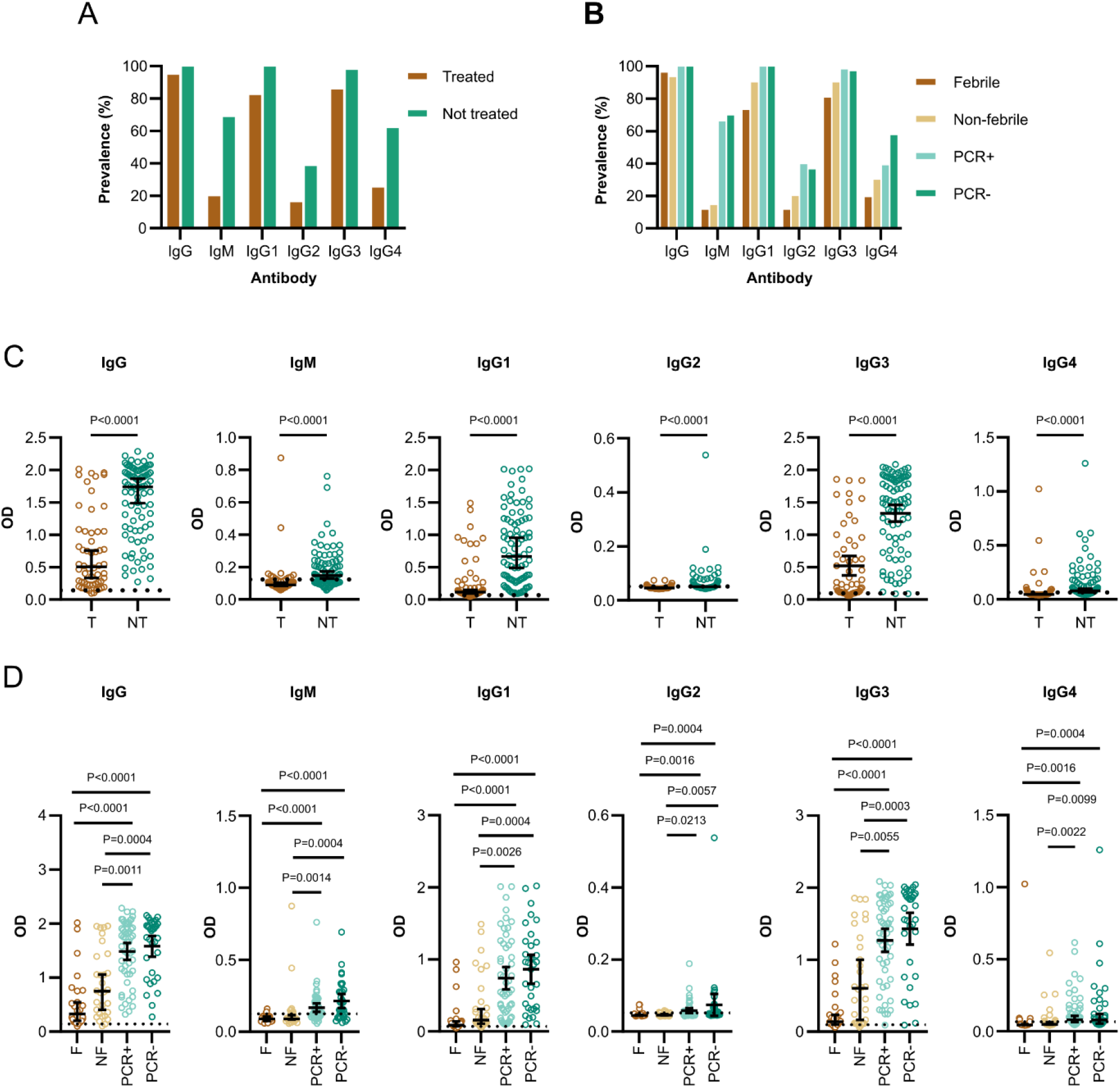
High antibody levels against MSP1_FL_ in volunteers who were protected from sporozoite challenge. (**A**) The prevalence of IgG, IgM and IgG subclass 1-4 antibodies in treated (T, n=56) and non-treated volunteers (NT, n=86). (**B**) The prevalence of IgG, IgM and IgG subclass 1-4 antibodies compared between the subgroups based on parasite growth patterns, treated febrile (F, n=26), treated non-febrile (NF, n=30), non-treated PCR positive (PCR+, n=53) and non-treated PCR negative (PCR-, n=33). (**C**) Levels of IgG, IgM and IgG subclass 1-4 antibodies compared between treated and non-treated volunteers. (**D**) Levels of IgG, IgM and IgG subclass 1-4 antibodies were compared between the four different subgroups. Each data point represents antibody levels for one sample in duplicates. The seropositivity cut-off value was calculated as the optical density (OD) of malaria-naïve plasma samples plus three standard deviations indicated as the dotted line. Error bars represent the median plus 95% confidence intervals. Statistical differences between treatment outcomes were calculated using Mann-Whitney test and between subgroups using Kruskal Wallis test with Dunn’s multiple comparisons test.

We investigated IgG subclasses and found that the response was dominated by cytophilic IgG1 (82%:100%, treated versus non-treated) and IgG3 (86%:98%, treated versus non-treated), while non-cytophilic IgG2 (16%-38%, treated versus non-treated) and IgG4 (25%:62%, treated versus non-treated) antibodies were less abundant (**Fig. 1A**). Within clinical subgroups, non-treated volunteers that were either PCR+ or PCR-tended to have a higher prevalence of anti-MSP1_FL_ antibodies compared to those that were treated and either febrile or non-febrile. However, the overall difference for total IgG and cytophilic IgG between subgroups was small. (**Fig. 1B**).

In marked contrast to the antibody prevalence, anti-MSP1_FL_ IgG, IgM and IgG subclass antibody levels were significantly higher in non-treated versus treated volunteers (*p*<0.0001, **Fig. 1C**). Within subgroups, both the non-treated PCR+ and PCR-volunteers had significantly higher anti-MSP1_FL_ antibody levels than those that were treated and either febrile or non-febrile. (*p*<0.0001-0.0213, **Fig. 1D**).

Next, we compared the magnitude of the responses between antibody isotypes and observed that IgG levels were significantly higher (*p*<0.0001) than IgM. Similarly, the levels of cytophilic IgG1 and IgG3 subclass antibodies were similarly higher (*p*<0.0001) than the non-cytophilic IgG2 and IgG4 (**Fig S1**); however, caution should be taken when comparing the magnitude of responses between the antibody classes due to potential differences in the sensitivity of secondary antibodies.

We observed significant and positive correlations between antibody levels against MSP1_FL_ and merozoites (**Fig S2**) suggesting co acquisition of anti-MSP1_FL_ antibodies alongside antibodies against other merozoite antigens. The strongest correlations were observed for cytophilic IgG1 (*r*=0.81, CI 0.75 – 0.86, *p*<0.0001) and IgG3 (r=0.82, CI 0,76 to 0,87, *p*<0.0001) while for non-cytophilic IgG2 and IgG4 antibodies only moderate correlations were detected (r=0.46-0.59, *p*<0.0001).

### IgG antibodies detect each subunit of MSP1_FL_ and are cross-reactive

We next investigated whether the IgG response was preferentially directed towards specific regions of the MSP1 molecule. We compared responses against the p83, p30, p38 and p42 subunits of MSP1 in non-treated and treated CHMI volunteers. As shown in **Figure 2A**, we observed antibody responses directed against all MSP1 subunits and found that non-treated volunteers had higher prevalence of antibodies compared to treated volunteers. This was observed for p83 (61%:91%, treated versus non-treated), p38 (23%:63%, treated versus non-treated) and p42 (59%:67%, treated versus non-treated). For p30, only 16% of the non-treated and 23% of the treated volunteers showed detectable antibody responses.

**Figure 2:**
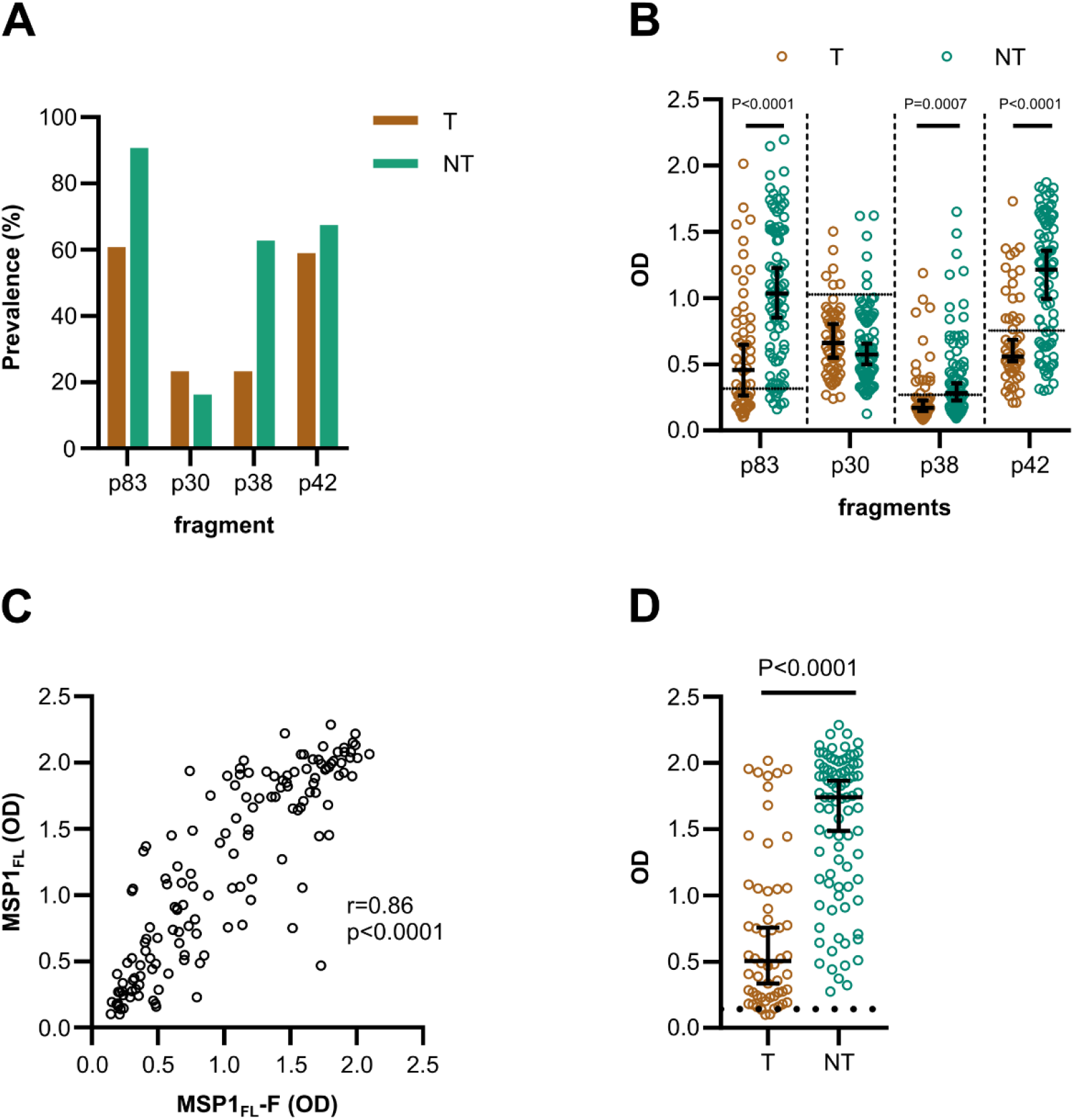
Antibody levels against subunits were high in non-treated volunteers and cross-reactive. (**A**) The prevalence of IgG antibodies in treated (T, n=56) and non-treated volunteers (NT, n=86). (**B**) IgG antibody levels against MSP1 subunits were compared between treated (T, n=56) and non-treated volunteers (NT, n=86). (**C**) Spearman’s correlation of anti-MSP1_FL_ (3D7) and MSP1_FL_-F (K1) antibody levels (OD) for CHMI volunteers (n=142). (**D**) IgG antibody levels against MSP1_FL_-F were compared between treated (T, n=56) and non-treated volunteers (NT, n=86). Each data point represents antibody levels for one sample in duplicates. The seropositivity cut-off value was calculated as the optical density (OD) of malaria-naïve plasma samples plus three standard deviations indicated as the dotted line. Error bars represent the median plus 95% confidence intervals. Statistical differences between treatment outcomes were calculated using Mann-Whitney test

In line with the previously reported antibody responses against full-length MSP1. The antibody levels against p83, p38 and p42 were significantly higher (*p*<0.0001-0.0007, **Fig. 2B**) for non-treated volunteers compared to treated individuals. However, for p30, no significant difference between the groups was observed. Importantly to note, for p30 and p42 we also observed higher background signals (OD=0.7-1.0) compared to p38 and p83 (OD=0.3), potentially due to antibody cross-reactivities from malaria naïve adults. Despite the different background intensities, the immunogenicity profiles are comparable with previous results from semi-immune adults living in Burkina Faso (Woehlbier et al., 2006).

MSP1 exists in two main allelic forms, represented by the MAD20 and K1 variant (Tanabe et al., 1987). Our ELISA assays utilized MSP1_FL_ based on the *P. falciparum* 3D7 strain that is like the MAD20 variant at this locus. We tested whether the antibodies were cross-reactive with MSP1_FL_-F that is based on the K1 variant. We found a high correlation between both variants (*r*=0.86, 95% CI 0.81-0.9, *p*<0.0001 [Spearman’s rho], **Fig. 2C**). Not surprisingly therefore, non-treated volunteers had significantly higher IgG levels against MSP1_FL_-F compared to treated volunteers (*p*<0.0001, **Fig. 2D)** which implies that antibodies bind to conserved and/or dimorphic regions.

### IgG from non-treated volunteers bind to conserved and dimorphic epitopes across the MSP1_FL_ molecule

To better localize antibody binding, we mapped linear B-cell epitopes using an MSP1_FL_ (3D7) peptide chip consisting of 1,715 15-mer peptides with a peptide-peptide overlap of 14 amino acids. We randomly selected 10 plasma samples from each clinical subgroup and analysed their linear epitope repertoire. As shown in **Figure 3**, we identified numerous epitopes across the entire MSP1_FL_ molecule that induce an IgG response. Dominant epitopes were found in the conserved N- and C-terminal regions (EEITTK, position 56-61, p83 and SPLKTLSEVSIQTE, position 1150-1163, p38) and central dimorphic domains (ETEETEET, position 747-754, p30). The majority of treated febrile and treated non-febrile volunteers showed only little binding events and non-treated PCR+ and PCR-individuals showed significantly stronger responses to several conserved and dimorphic regions compared to treated volunteers (**Supplementary Table 1**).

**Figure 3:**
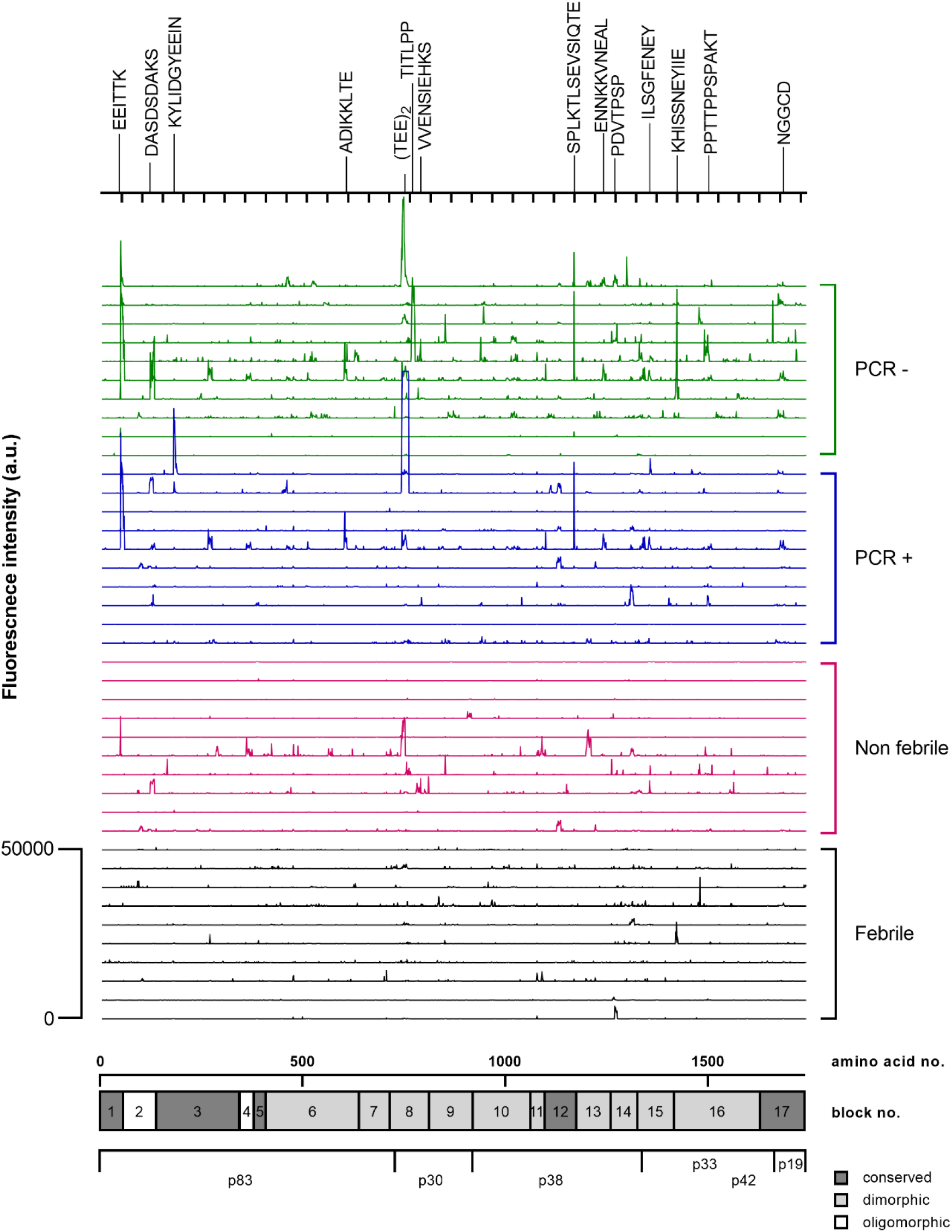
IgG from treated volunteers bind to conserved and dimorphic epitopes across the whole MSP1 molecule. Fluorescence intensity landscapes across MSP1_FL_ shown for individuals of the four subgroups based on parasite growth patterns: treated-febrile (n=10), treated non-febrile (n=10), non-treated PCR positive (n=10) and non-treated PCR negative (n=10). Every line represents a sample. Relevant epitopes have been highlighted on top. A graphical representation of the primary structure of MSP1_FL_ is shown below the fluorescence intensity landscapes.

### Anti-MSP1_FL_ antibodies induce high levels of Fc-mediated effector functions that are associated with protection

We next investigated whether MSP1_FL_ was a target of opsonizing antibodies that could induce Fc-mediated effector mechanisms. Therefore, we used four antigen-specific *in vitro* functional assays to measure antibody-dependent complement fixation (AbC’) (Boyle et al., 2015; Reiling et al., 2019), opsonic phagocytosis activity (OPA) of MSP1_FL_-coupled fluorescent beads by monocytes (Kana et al., 2019), antibody-dependant respiratory burst (ADRB) of neutrophils (Joos et al., 2015) and antibody-dependent natural killer cell (Ab-NK) activities (Odera et al., 2021). The latter was assessed by multiparameter flowcytometry resulting in two readouts: degranulation of NK cells (Ab-NK:CD107a) and IFNγ production (Ab-NK:IFNγ).

We compared MSP1_FL_-specific Fc-mediated effector functions between volunteers who required treatment and those who did not. We found a higher prevalence of functional antibodies across all five effector mechanisms for non-treated compared to treated volunteers (**Fig. 4A, B**). Similarly, the magnitude of Fc-mediated functions was significantly higher for non-treated volunteers compared to those who were treated (AbC’; *p*<0.0001, OPA; *p*<0.0001, ADRB; *p*<0.0001; Ab-NK:CD107a; *p*=0.0175, Ab-NK:IFNγ; *p*=0.0203, **Fig. 4C**). We also observed that non-treated PCR+ and PCR-individuals had significantly higher AbC’, OPA and ADRB activity compared to treated febrile or non-febrile individuals. For Ab-NK activities, we only observed significant differences in degranulation (Ab-NK:CD107a) for non-treated PCR+/-versus treated febrile individuals but IFNγ production (Ab-NK:IFNγ) was not significantly different across the subgroups (**Fig. 4D**).

**Figure 4:**
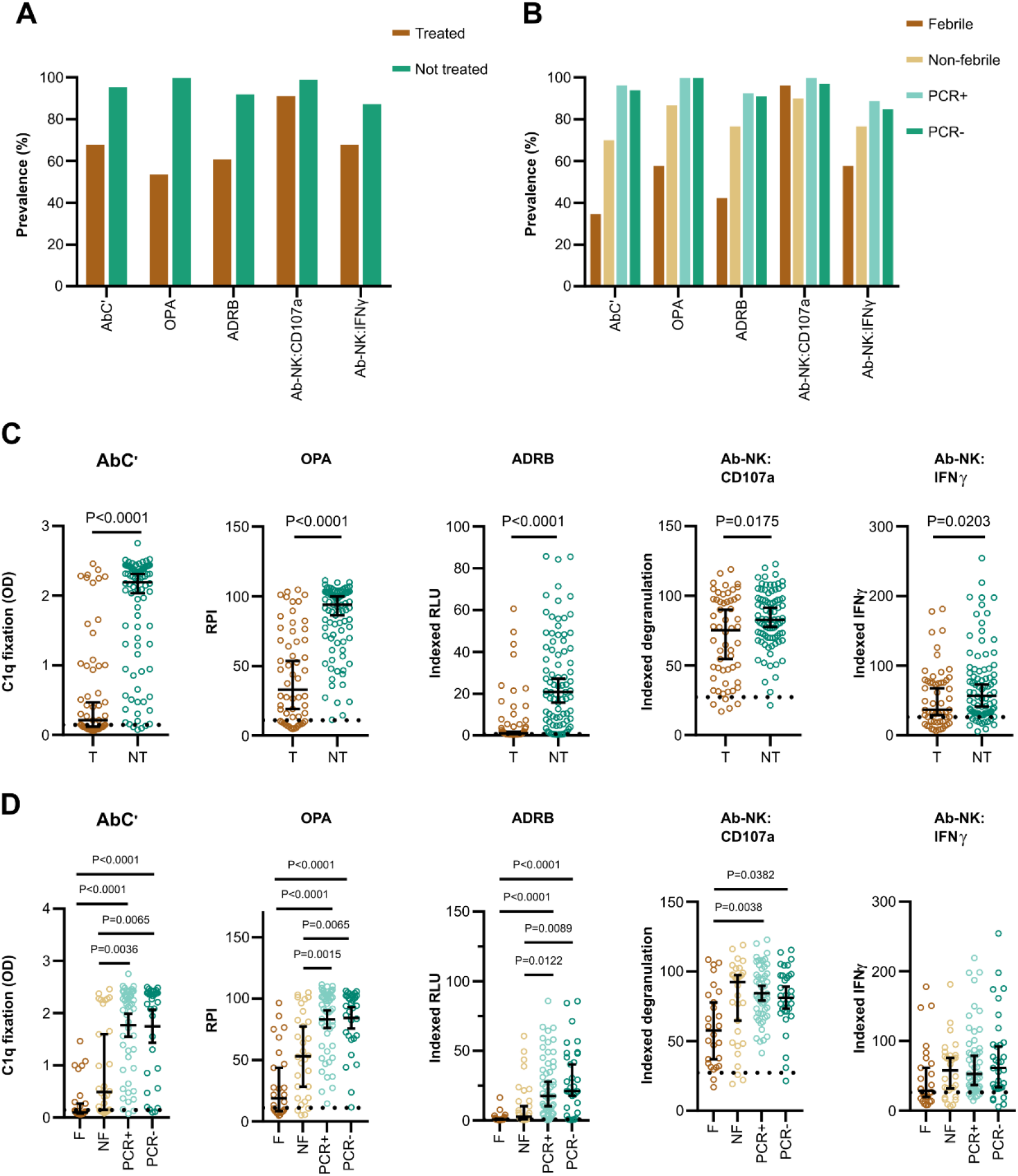
Prevalence and magnitude of MSP1_FL_-specific Fc-effector functions are high in non-treated volunteers. (**A**) The prevalence of MSP1_FL_ -specific Fc-mediated functional activities in treated (T, n=56) and non-treated volunteers (NT, n=86). (**B**) The prevalence of MSP1_FL_ -specific Fc-mediated functional activities compared between the subgroups based on parasite growth patterns, treated febrile (F, n=26), treated non-febrile (NF, n=30), untreated PCR positive (PCR+, n=53) and untreated PCR negative (PCR-, n=33). (**C**) Levels of Fc-mediated effector functions of anti-MSP1_FL_ antibodies were compared between treated and non-treated volunteers. (**D**) Levels of Fc-mediated effector functions of anti-MSP1_FL_ antibodies were compared between the different subgroups, based on parasite growth densities: treated febrile, treated non-febrile, non-treated PCR positive and non-treated PCR negative. Each data point represents antibody levels for one sample in duplicates. The seropositivity cut-off value was calculated as the activity level of malaria-naïve plasma samples plus three standard deviations indicated as the dotted line. Error bars represent the median plus 95% confidence intervals. Statistical differences between treatment outcomes were calculated using Mann-Whitney test and between subgroups using Kruskal Wallis test with Dunn’s multiple comparisons test. CD107a; Fc-mediated natural killer cell degranulation, IFNγ; Fc-mediated natural killer IFNγ production, ADRB; antibody-dependent respiratory burst by neutrophils, OPA; opsonic phagocytosis activity of MSP1_FL_-coupled microsphere beads by monocytes, AbC’; antibody-dependent complement fixation activity.

We further explored the potential relationship between Fc-mediated effector functions and antibody levels and observed high correlations between anti-MSP1_FL_ IgG and AbC’ (*r*=0.83, 95% CI 0.77-0.88, *p*<0.0001), ADRB (*r*=0.91, 95% CI 0.88-0.94, *p*<0.0001) and OPA (*r*=0.89, 95% CI 0.85-0.92, *p*<0.0001, **Fig. 5A)**. For both readouts of the Ab-NK assay, we observed moderate (*r*>0.5) correlations with IgG. Correlations with IgM, were low with Ab-NK (r>0.2), and moderate (*r*>0.5) with AbC’ (*r*=0.58, 95% CI 0.46-0.69, *p*<0.0001), ADRB (*r*=0.60, 95% CI 0.48-0.70, *p*<0.0001) and OPA (*r*=0.55, 95% CI 0.42-0.66, *p*<0.0001). Cytophilic antibodies (IgG1 and IgG3) can efficiently bind complement and most of the Fc-receptors (FcRs) of immune cells (Vidarsson et al., 2014). As expected, the correlations between cytophilic antibodies and effector functions were higher compared to non-cytophilic antibodies (*r*=0.43-0.90 vs. 0.29-0.66, respectively). Next, the correlations between effector functions were explored (**Fig. 5A**). OPA, ADRB and AbC’ were highly correlated with each other (*r*=0.86-0.92, *p*<0.0001) but less so for Ab-NK (*r*=0.47-0.59, *p*<0.0001). As expected, the two readouts of the Ab-NK assay (CD107a and IFNγ) were highly correlated (*r*=0.88, 95% CI 0.84-0.91, *p*<0.0001) with each other.

**Figure 5:**
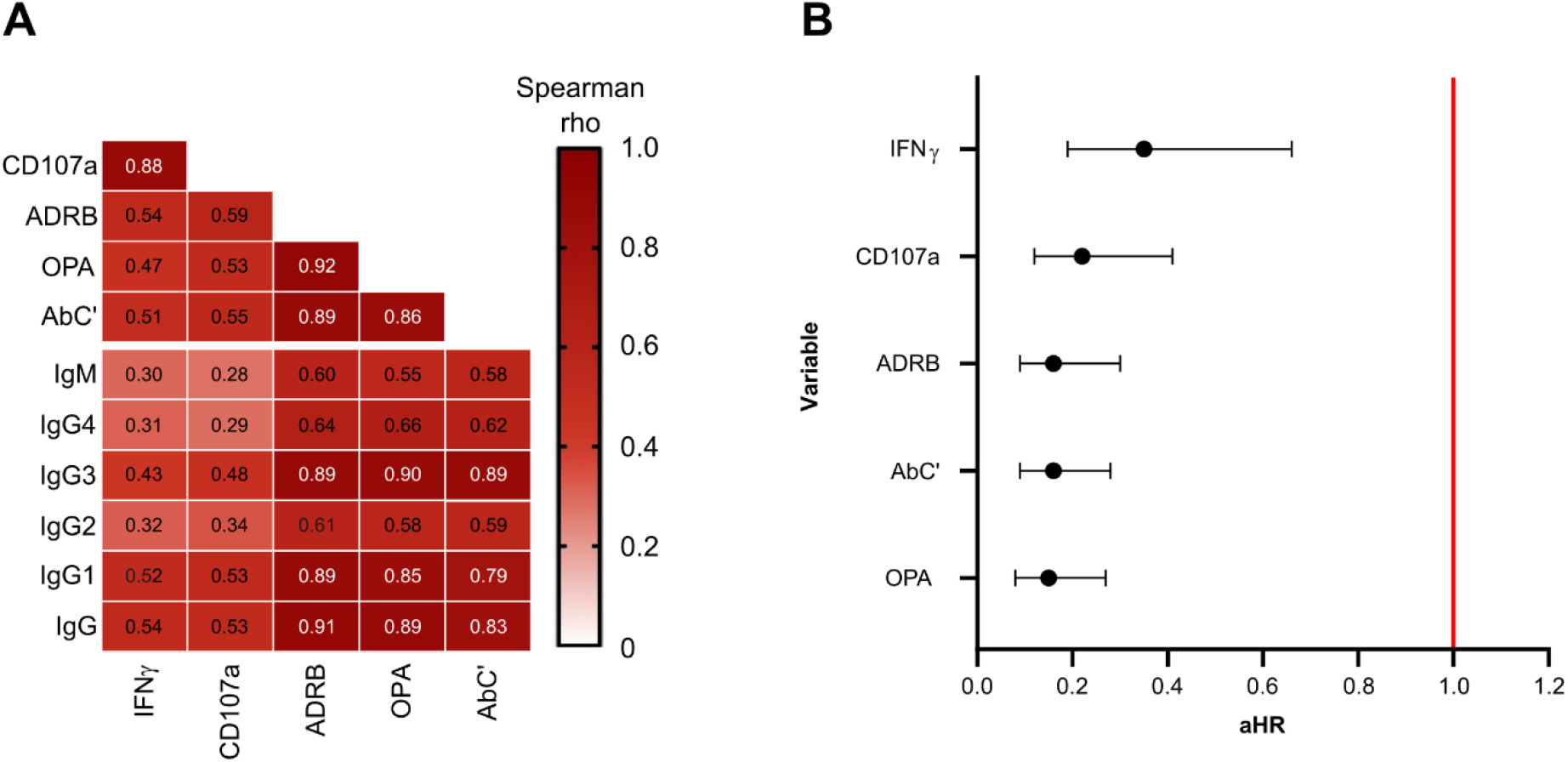
MSP1_FL_-specific functional activities are correlated with each other and antibody levels and associated with protection. (**A**) A heatmap with correlation matrix showing spearman rank correlation coefficients of antibody levels and Fc-mediated effector functions. The color intensity represents the strength of correlation. (**B**) Forest plot showing adjusted hazard ratios (aHR) for MSP1_FL_-specific functional activities ranked from lowest to highest. The aHRs were calculated using the cox regression model comparing time to diagnosis between high versus low responders based on function-specific thresholds while adjusting for confounders (drug levels and year of study). Error bars indicate 95% confidence intervals and the red line indicates no protection (aHR = 1.0). CD107a; Fc-mediated natural killer cell degranulation, IFNγ; Fc-mediated natural killer IFNγ production, ADRB; antibody-dependent respiratory burst by neutrophils, OPA; opsonic phagocytosis activity of MSP1_FL_-coupled microsphere beads by monocytes, AbC’; antibody-dependent complement fixation activity.

To identify MSP1_FL_-specific effector functions that may be important for protection against malaria in the CHMI study, we analysed the association of each effector function individually with the time to treatment during follow up. To do this, responses were converted into two categories (high and low) based on function-specific thresholds using maximally selected rank statistics (Musasia et al., 2022; Nkumama et al., 2022; Odera et al., 2021). Cox proportional hazards were adjusted for residual lumefantrine levels and year of study as confounders. Interestingly, we observed statistically significant and strong associations with protection for each MSP1_FL_-specific effector function with aHR estimates ranging between 0.15 to 0.35 (**Fig. 5B**).

### The breadth of MSP1_FL_-specific effector functions is a strong predictor of protection from malaria

Since we showed that each MSP1_FL_-specific Fc-effector function predicted significantly reduced risk of requiring treatment upon sporozoite challenge, we next wanted to assess the contribution of the breadth of MSP1_FL_-specifc function to protective immunity. Therefore, we developed breadth scores of functional activities for every volunteer. We categorized the level of Fc-mediated functions of the study participants as either high or low (coded 1 or 0, respectively) based on function specific thresholds. We then summed the breadth scores across the five functions (Nkumama et al., 2022).

Non-treated volunteers had higher breadth scores compared to treated volunteers, with 78% (67/86) versus 29% (16/56), respectively (**Fig. 6A**). Furthermore, using a Kaplan-Meier survival analysis, we showed that protection increased stepwise with rising breadth scores. A breadth score of zero was only associated with a protective efficacy of 7% compared to 81% for that of five (*p*<0.0001, **Fig. 6B**). This outcome, highlights that the breadth of MSP1_FL_-specific functions is a stronger correlate of protection than individual functions.

**Figure 6:**
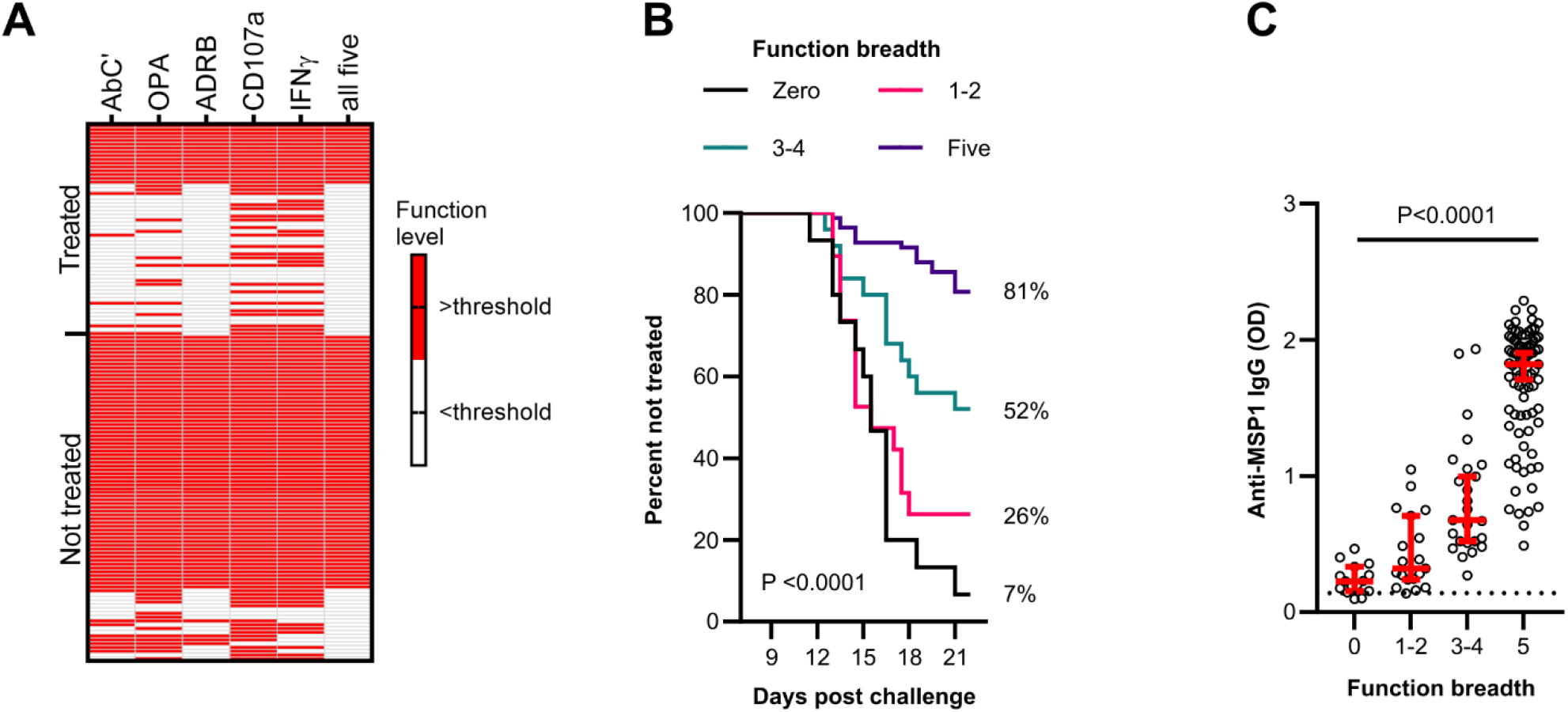
Breadth of MSP1_FL_-specific effector functions is a strong predictor of protection from malaria. (**A**) A heatmap showing the activity levels of all five Fc-mediated effector functions in treated (n=56) and non-treated (n=86) volunteers. Responses above a function-specific threshold are shown in red. Each column is a Fc-mediated function while each row is a single volunteer. AbC’; antibody-dependent complement fixation activity, OPA; opsonic phagocytosis activity of MSP1_FL_-coupled microsphere beads by monocytes, ADRB; antibody-dependent respiratory burst by neutrophils, IFNγ; Fc-mediated natural killer IFNγ production, CD107a; Fc-mediated natural killer cell degranulation. (**B**) Kaplan-Meier plot of volunteers who remained non-treated at different timepoints over the course of 21 days post challenge. Each line represents a function breadth score starting from 0 (n=15), 1-2 (n=19), 3-4 (n=25) and 5 (n=83). Significance was assessed using Log rank sum test. (**C**) Anti-MSP1_FL_ IgG levels were compared between individuals with different levels of MSP1_FL_-specific breadth of function. Each dot represents one sample in duplicates. Error bars represent the median plus 95% confidence intervals. The seropositivity cut-off value was calculated as the optical density (OD) of malaria-naïve plasma samples plus three standard deviations indicated as the dotted line. Statistical differences were calculated using Kruskal-Wallis test.

Moreover, we observed that the breadth of function increased with rising levels of anti-MSP1_FL_ IgG (**Fig. 6C**), highlighting the importance of high anti-MSP1_FL_ antibody titers for protection.

## Discussion

MSP1 has been extensively analysed in the context of naturally-acquired and vaccine-induced immunity. However, most studies focused on the conserved C-terminal subunit representing ∼20% of the molecule (Ellis et al., 2010; Fowkes et al., 2010; Malkin et al., 2007; Ogutu et al., 2009; Richards et al., 2013). Furthermore, although the functional immunological correlate for these and other studies has widely been invasion-inhibition, recent data call this into question in favour of Fc-mediated effector mechanisms (Nkumama et al., 2022; Reiling et al., 2019). Our new data now show that the entire full-length antigen is immunogenic and that these antibodies are strong inducers of Fc-mediated effector functions, which in turn are strongly associated with protection from malaria. Taken together, these data suggest that previous investigations not only missed important epitopes within MSP1, but also assessed its’ functional relevance sub-optimally.

The finding that the entire MSP1 molecule was the target of functional antibodies was not surprising and is supported by early and more recent studies (Jäschke et al., 2017; Woehlbier et al., 2006). The most striking results were that anti-MSP1_FL_ antibodies almost completely recapitulated our recent data identifying Fc-mediated mechanisms targeting merozoites as strong correlates of protective immunity (Nkumama et al., 2022; Odera et al., 2021). These data are consistent with the fact that MSP1 is the most abundant protein on the surface of merozoites (Gilson et al., 2006). Although individual Fc-mediated mechanisms have been correlated with protection from malaria in unrelated studies (Boyle et al., 2015; Hill et al., 2013; Joos et al., 2010; Kana et al., 2019; Murungi et al., 2016; Osier et al., 2014; Tiendrebeogo et al., 2015), including those specifically induced by MSP1^19^ (Kana et al., 2019; Reiling et al., 2019), the breadth of MSP1_FL_-induced Fc-function across multiple effectors has not previously been evaluated in a single study. Remarkably, as observed with IgG against merozoites (Nkumama et al., 2022), the breadth of anti-MSP1_FL_ IgG Fc-mediated function was the strongest predictor of protection, suggesting it could contribute to a high level of immunity as a monovalent subunit vaccine.

We found that anti-MSP1_FL_ IgG antibodies were predominantly comprised of the cytophilic IgG1 and IgG3 isotypes. This is in keeping with data from multiple studies involving both the N- and C-termini of MSP1_FL_ (Cavanagh et al., 2001; Cavanagh et al., 2004; Egan et al., 1995; Stanisic et al., 2009), with our own data using whole merozoites (**Fig S2**) and supports the well-established role for cytophilic antibodies in promoting immune effector activity that could be detrimental to parasites. Interestingly, a proportion of individuals had cytophilic anti-MSP1_FL_ antibodies that induced Fc-mediated function across multiple effectors but were not protected from CHMI. An analysis of the correlation between these different effector functions revealed high correlations between complement activity, monocyte phagocytosis and neutrophil respiratory burst, but these were slightly lower for natural killer cell degranulation and IFNγ secretion. Taken together with the data showing that the breadth of effector function was the strongest correlate of protection, this may suggest that distinct epitopes within MSP1_FL_ induce the full range of immune effectors. Similar observations have been made for the circumsporozoite surface protein (CSP), the major surface antigen of *Plasmodium falciparum* sporozoites. While all regions of the protein were targets of opsonizing antibodies inducing phagocytosis of sporozoites, antibodies targeting the N-terminal domain in particular were most effective in activating phagocytes (Feng et al., 2021).

Alternatively, such antibodies may target similar epitopes but differ quantitatively or qualitatively. For example, the glycosylation status of IgG antibodies has been shown to be critical for receptor engagement and induction of NK cell degranulation which played an important immunological role in malaria (Larsen et al., 2021b) and other infectious diseases such as COVID-19 (Larsen et al., 2021a). Additional possibilities are that FcγR polymorphisms account for the variation in protection despite the presence of anti-MSP1_FL_ antibodies, or indeed that antibodies to other antigens are more important for a protective immune response in some individuals. Additional investigations in this regard will be necessary.

We were surprised to detect IgM antibodies against MSP1_FL_ in malaria-exposed adults that presumably had life-long exposure to malaria parasites. Although we showed that antibody responses against MSP1_FL_ were dominated by cytophilic IgG, recent findings have shown that IgM persists over time, activates complement-mediated invasion inhibition of red blood cells and is associated with protection from malaria (Boyle et al., 2019). Additionally, it was demonstrated that IgM from malaria exposed adults induces merozoite phagocytosis by monocytes (Hopp et al., 2021). However, research of FcμR distribution among innate immune cells such as macrophages has led to conflicting results (Honjo et al., 2013; Hopp et al., 2021; Kubagawa et al., 2009; Lang et al., 2013; Shima et al., 2010; Uher et al., 1981) in part due to differences in activation status of the cells and thus expression levels of FcμR. In this study we did not purify IgG or IgM because of limited sample volumes and thus could not assess its’ specific functional activity. Nevertheless, we found that the correlations between IgM antibody levels and IgG effector functions were relatively low. Additionally, while the data on IgG and IgM cannot be directly compared due to differences in the quality of secondary reagents, it is tempting to speculate that the IgM findings were a red herring, as the antibody levels were relatively low compared to IgG. We cautiously speculate that MSP1_FL_-specific IgM plays a minor role in protection from malaria amongst semi-immune adults.

IgG antibodies between the two main allelic variants of MSP1_FL_ were highly correlated suggesting the presence of cross-reactive or shared epitopes and bodes well for vaccine development. Additional analyses using linear overlapping peptides confirmed not only that B-cell epitopes were present throughout the entire length of MSP1_FL_, but also showed that in contrast to treated volunteers, IgG antibodies from those that did not require treatment preferentially targeted epitopes located within conserved and dimorphic regions. We did not have sufficient reagents to test whether these specific peptides induced functional antibodies in our panel of Fc-dependent assays. Importantly, these peptides are distributed across the entire MSP1 molecule and supports our view that the full-length antigen should be prioritised for vaccine design. Indeed, a recent phase 1 clinical trial with full-length MSP1 (SumayaVAC-1) was safe and induced high titers of strain-transcending antibodies with complement fixation and ADRB activity in malaria-naïve adults (Blank et al., 2020).

We found that breadth of MSP1_FL_-specific functional activity was low in the majority of treated volunteers; however, the fact that 29% of volunteers in whom the breadth of function was high were not protected from malaria, was not surprising. Although MSP1_FL_ is the most abundant antigen on the merozoite surface and therefore might dominate an antibody-mediated protective immune response against merozoites, other and/or multiple antigens could be important for protection which has been highlighted in previous studies (Kana et al., 2019; Osier et al., 2008; Reiling et al., 2019; Richards et al., 2013). Moreover, we and others have recently shown that antibodies against antigens from other blood stages such as variant specific surface antigens (VSA) on trophozoites (Kimingi et al., 2022) and ring stage antigens (Musasia et al., 2022) were significantly associated with a reduced risk of developing clinical malaria in CHMI volunteers. As such, it remains plausible that antibodies targeting several blood stage antigens might work in synergy to confer protection. It is also possible that other antibody functions that were not measured as part of this investigation are important. These could include neutrophil-mediated phagocytosis, antibody-dependent cellular inhibition by monocytes (ADCI) (Bouharoun-Tayoun et al., 1995) or ADCC by γδ T cells (Farrington et al., 2020) amongst others.

Our study had several limitations. We were unable to assess the functional activity of antibodies directed against individual subunits and specific peptides within MSP1. Furthermore, our MSP1 protein reagents were expressed using different systems and limit our interpretations of some of the data due to high background signals. The data on linear B-cell epitopes likely misses important conformational epitopes that we identified in a previous study (Kamuyu et al., 2018).

These limitations notwithstanding, we show that MSP1_FL_ recapitulate the breadth of Fc-mediated functional activity targeting merozoites and could account for over 80% of clinical protection against malaria in the CHMI study. These data raise optimism that malaria vaccines designed to interrupt transmission at the blood stage of the parasite life-cycle should incorporate full-length MSP1.

## Materials and methods

### Study design and samples

The design of the Controlled Human Malaria Infection of Semi-Immune Kenyan Adults (CHMI-SIKA) study has been previously described (Kapulu et al., 2018). Briefly, 161 healthy Kenyan adults aged 18-45 years with different antibody levels against schizont extract were recruited from three consecutive cohorts between 2016 and 2018. The volunteers were challenged with 3200 cryopreserved *Plasmodium falciparum* NF54 sporozoites (Sanaria) by direct venous inoculation (DVI). Testing for blood stage parasitemia was conducted by qPCR twice per day between day 7 and day 14 and once per day between day 15 and day 21 post challenge. Volunteers were treated with artemether-lumefantrine when blood stage levels exceeded 500 *P*.*f*./μl, symptoms of clinical malaria with detectable parasites were recorded or after follow-up at day 22.

Data from 19 volunteers were excluded due to antimalarial drug levels above the reported minimum inhibitory concentration in 12 volunteers and the presence of non-NF54 parasites in 7 volunteers. Thus, plasma samples from 142 volunteers (31 % female) collected one day before challenge (C-1) were used in this study.

### Ethics statement

The CHMI study was conducted at the KEMRI Wellcome Trust Research Programme in Kilifi, Kenya with ethical approval from the KEMRI Scientific and Ethics Review Unit (KEMRI//SERU/CGMR-C/029/3190) and the University of Oxford Tropical Research Ethics Committee (OxTREC 2–16). All participants gave written informed consent. The study was registered on ClinicalTrials.gov (NCT02739763), conducted based on good clinical practice (GCP), and under the principles of the Declaration of Helsinki.

### Expression of recombinant merozoite proteins

The Plasmid containing the codon-optimized sequence of full-length MSP1 (3D7) was received from the plasmid repository Addgene (Plasmid #47709) which has been optimized for expression in mammalian cells (Crosnier et al., 2013; Kamuyu et al., 2018). MSP1_FL_ was expressed in the Expi293 expression system (Gibco) following the manufacturer’s instructions. In brief, 2 × 10^6^ cells/ml of Exp1293F culture were transfected with the expression plasmid using the ExpiFectamine 293 transfection kit (Gibco). At 20 hours post transfection, the transfection enhancers were added and after 5 days recombinant proteins were harvested and purified from culture supernatant using the Ni-NTA purification system (Invitrogen).

Recombinant MSP1-subunits and MSP1_FL_-F were expressed in *E*.*Coli* as previously described (Kauth et al., 2003; Kauth et al., 2006) and kindly provided by Sumaya Biotech.

### Indirect Enzyme-linked Immunosorbent Assay (ELISA)

Recombinant MSP1_FL_ and MSP1 subunits were coated with 0.5mg/well onto 96-well plates at 4°C overnight. The next day, the plates were washed with 1x phosphate buffered saline (PBS) containing 0.05% Tween 20 (PBST) and blocked with 200μl/well 1% skimmed milk for 2 hours at room temperature. After blocking, the plates were washed followed by incubation with 50μl/well serum samples at 1:1000 for 2 hours at room temperature. After incubation, plates were washed and 50μl/well of respective secondary antibodies conjugated with Horseradish peroxidase (HRP) were added for 1 hour at room temperature: rabbit anti-human IgG (Agilent), goat anti-human IgM (Thermo Fisher), and rabbit anti-human IgG1, 2, 3 or 4 (The Binding Site). Afterwards, the plates were washed and substrate solution (0.4mg/ml O-phenylenediamine (OPD), Sigma-Aldrich) was added and incubated in the dark at room temperature for 30 min. The reaction was stopped by adding 15μl/well of 1M hydrochloric acid (HCL) and the absorbance was read at 492nm using the BioTek Cytation 3 cell imaging multi-mode reader and the Gen5 v3.02 software.

### Mapping of linear B-cell epitopes

The mapping of linear IgG epitopes against full-length MSP1 was performed by PEPperPRINT GmbH, Heidelberg as previously described (Blank et al., 2020). Briefly, the sequence of full-length MSP1 (3D7) (UniProt ID: Pf3D7_0930300) was elongated with neutral GSGSGSG linkers at the N- and C-terminus to avoid truncated proteins. Next, the modified protein sequence was converted into short 15-mer amino acid peptides with a 14 amino acid overlap between the peptides. Subsequently, 1,720 unique peptides were printed in duplicates on the chip framed by additional HA (YPYDVPDYAG, 62 spots) and polio (KEVPALTAVETGAT, 62 spots) control peptides. The chip was blocked with Rockland blocking buffer MB-070 for 30 min, followed by pre-staining with secondary goat anti-human IgG (Fc) DyLight680 (0.1 μg/ml) and the monoclonal mouse anti-HA control antibody (12CA5) DyLight800 (0.5 μg/ml in incubation buffer (washing buffer with 10% blocking buffer) for 45 min. Next, other copies of the MSP1_FL_ microarray were incubated with 40 randomly selected human plasma samples (10 per clinical subgroup) from the CHMI-SIKA study (1.1000 in incubation buffer) for 16 h at 4C while shaking at 140 rpm. After washing with PBST, fluorescently labelled secondary antibodies were added for 45 min at room temperature. The signals were detected using an InnoScan 710-IR Microarray Scanner at scanning gains of 50/50 (red/green). Spot intensity quantification was based on 16-bit grey scale tiff files with higher dynamic range than the 24-bit colourized tiff files. The analysis of the data was done with PepSlide Analyzer. A maximum spot-to-spot deviation of 40% was tolerated, otherwise the intensity value was put to zero.

### Antibody-dependent complement fixation (AbC’) assay

AbC’ activity of MSP1_FL_-specific antibodies was assessed in a modified ELISA measuring fixation of C1q, the first component of the classical complement pathway using a published protocol (Boyle et al., 2015; Reiling et al., 2019). Briefly, 96-well plates (Thermo Fisher) were coated with 0.5mg/well of MSP1_FL_ overnight at 4°C. The following day, plates were washed four times with PBST and blocked with 200μl/well of 1% Casein/PBS at 37°C for 2 h. After blocking, the plates were washed and 50μl/well of plasma samples diluted at 1:10 in PBS were added for 2 h at 37°C. Following incubation, the plates were washed and 40μl of recombinant C1q (Abcam) at 10mg/ml diluted in blocking buffer was added for 30 min at 37°C. Thereafter, the plate was washed and incubated with 50μl/well of sheep anti-human C1q-HRP (Abcam) at a 1:100 dilution in blocking buffer for 1 h at 37°C. After washing, 50μl/well of OPD solution (Sigma-Aldrich) was added and incubated for 45 min at room temperature before the reaction was stopped by adding 15μl/well of 1M HCL. The absorbance was read at 492nm using the BioTek Cytation 3 cell imaging multi-mode reader and the Gen5 v3.02 software.

### Antibody-dependent respiratory burst (ADRB) assay

The ADRB assay was performed as previously described (Kapelski et al., 2014; Nkumama et al., 2022). Briefly, MSP1_FL_ at 0.5mg/well was coated onto opaque 96-well plates overnight (Greiner) at 4°C. The following day, the plates were washed with sterile PBS and blocked with 200μl/well of sterile 1% casein/PBS for 1 hour at 37°C. After blocking, the plates were washed and incubated with 50μl/well of plasma samples diluted in PBS at 1:10 for 1 hour at 37°C.

Polymorphonuclear leucocytes (PMN) were prepared from whole blood collected in Heparin vacutainers. Blood was diluted 1:1 with hanks balanced salt solution (HBSS, Thermo Fisher), carefully layered on top upon 7 ml of Histopaque^®^-1077 (Sigma-Aldrich) and centrifuged at 600 x g for 15 min. The pellet containing the PMN was resuspended in HBSS, mixed with 3% Dextran in a 1:2 ratio and incubated for 1 hour at room temperature. Next, the supernatant was collected and centrifuged at 500 x g for 7 min at 4°C. The pellet was then resuspended in ice cold 0.2% NaCl for 30 seconds to lyse contaminating RBC followed by adding an equal volume of ice cold 1.6% NaCl to stop lysis. Afterwards, the cells were centrifuged and the PMN were resuspended in PMN buffer (0.1% bovine serum albumin (BSA), 1% D-Glucose in HBSS) and counted using a hemocytometer. The concentration of viable neutrophils was adjusted to 10 × 10^6^ cells/ml and the cells were kept on ice.

Next, the plates were washed and 50μl/well of luminol (Sigma) at 0.04mg/ml was added prior to adding 50μl/well of neutrophils. Chemiluminescence at 450nm was immediately read for every 2 min over a duration 1.5 h using the Biotek Synergy 4 plate reader and the Gen 5 acquisition software. The maximal relative light unit (RLU) for each sample was generated and indexed based on responses of a pool of hyper immune sera from Kenyan adults (PHIS), the positive control. The ADRB index was calculated as: (RLU of samples) / (RLU of PHIS) x 100.

### Opsonic phagocytosis activity (OPA) assay

The opsonic phagocytosis assay of antigen-coupled beads was based on a published protocol (Kana et al., 2019). Briefly, polychromatic red 1μm microsphere beads (Polysciences Inc) were coupled with 30mg of MSP1_FL_ in borate buffer (Polysciences Inc) overnight at room temperature in the dark while rotating. The following day, the beads were centrifuged and supernatant was carefully removed. Next, the beads were blocked thrice in blocking buffer (10mg/ml BSA in borate buffer) for 30 min while rotating and stored in PBS with 5% glycerol and 0.1% sodium azide at 4°C.

For opsonization, 50μl containing 7.5 × 10^6^ antigen-coupled beads were added to each well of 96-well U-bottomed plates followed by incubation with 50μl/well of heat-inactivated serum samples diluted at 1:2000 for 1 h at 37°C. After incubation, the plates were centrifuged at 2000 x g for 7 min and washed thrice with PBS. Beads were resuspended in 50μl of THP1 cell culture medium (2mM L-glutamine, 2mM HEPES, 10% FCS, 1% Penicillin-streptomycin in RPMI 1640 media) and 50,000 THP1 cells in 150μl were added to each well for 30 min at 37°C. Phagocytosis was arrested by centrifugation at 1200 rpm for 7 min at 4°C. Plates were washed with ice-cold FACS buffer (0.5% BSA and 2mM ethylenediaminetetraacetic acid (EDTA) in PBS) and subsequently fixed in 2% formaldehyde/PBS. Flow cytometry was used to quantify THP1 cells containing fluorescent beads in the PE channel on the FACS CantoII high-throughput system (BD biosciences). Data analysis was performed using FlowJo V10.

Phagocytosis activity for each sample was indexed against the positive control (PHIS). The phagocytosis index was calculated as: (% of stained THP1 cells opsonized with samples) / (% of stained THP1 cells opsonized with PHIS) x 100.

### Antibody-mediated natural killer cells activation (Ab-NK)

The Ab-NK assay was performed as previously described (Odera et al., 2021). In brief, 500μg of MSP1_FL_ was coated onto 96 well-culture plates overnight at 4°C. The following day, the plates were washed with PBS and blocked for 1 h with 1%Casein/PBS at 37°C. After blocking, the plates were washed and incubated with 50μl/well of serum samples (1:10) for 4 h at 37°C. NK cells were isolated from human blood samples from healthy malaria naïve donors. First, peripheral blood mononuclear cells (PBMCs) were harvested using density gradient separation, washed and resuspended in NK cell culture medium (RPMI 1640 media with 2mM L-glutamine supplemented with 10% FCS and 1% Penicillin-streptomycin). NK cells were isolated from PBMCs by negative isolation using the NK cell isolation kit (Miltenyi Biotec) as per manufacturer’s instructions. A mix containing 5 × 10^5^ freshly isolated NK cells, anti-human CD107a PE (1:70, BD biosciences), brefeldin A (1:200, Sigma-Aldrich) and monensin (1:200, Sigma-Aldrich) was added into each well and incubated for 18 h at 37°C. After stimulation, NK cells were carefully transferred into 96 well V-bottomed plates, centrifuged at 1500 rpm for 5 min at 4°C and washed with ice-cold FACS buffer (1% BSA, 0.1% sodium azide in PBS). NK cell viability was assessed by staining with 10 μl/well of fixable viability dye eFluor™520 (Thermo Fisher) for 10 min at 4°C. NK cell surface receptors were stained with 20μl/well of an antibody mix consisting of anti-CD56 APC (1:17, BD biosciences) and anti-CD3 PE-Cy5 (1:33, BD biosciences) for 30 min at 4°C in the dark. Next, NK cells were washed and fixed in 80μl/well of fixing solution (CellFIX, BD biosciences) for 10 min at 4°C and subsequently permeabilized in 80μl/well of permeabilization buffer (permwash, BD biosciences) for 10 min at 4°C. Intracellular IFNγ was detected by adding 30 μl/well of anti-IFNγ PE-Cy7 (BD biosciences) diluted at 1:33 in permeabilization buffer for 1 h at 4°C in the dark. After intracellular staining, the cells were washed thrice with 150μl/well of permeabilization buffer and finally resuspended in 150μl/well FACS buffer. Acquisition was done on the FACS CantoII (BD biosciences) and the data was analysed using FloJo V10. NK cell activity (degranulation and IFNγ expression) for each sample was indexed against the positive control (PHIS). The degranulation/IFNγ index was calculated as: (% of NK cell degranulation/IFNγ of samples) / (% of NK cell degranulation/IFNγ release of PHIS) x 100.

### Statistical Analysis

Data was analysed using Prism 9.3.1 (GraphPad), Stata (version 14) or R. The Mann Whitney U test was used to compare medians between treated and non-treated groups. The Kruskal-Wallis test was used to compare the four phenotypes based on parasite growth densities (febrile, non-febrile, PCR+ and PCR-) combined by Dunn’s test for multiple comparisons. Pairwise correlations were calculated using nonparametric Spearman’s correlations. The Wilcoxon test combined with Hommel correction was used to identify epitopes that were significantly different between non-treated and treated volunteers. Functional activity levels were categorized in high and low responses based on function specific thresholds which were determined by using maximally selected rank statistics analysis method in R (Nkumama et al., 2022). The categorized data was analysed to assess the association between breadth of functional activity and time to treatment using the Cox proportional hazards model. Potential confounders were fit to the model (cohort and anti-malarial drug levels). The Log rank sum test was used to compare the Kaplan-Meier survival curves.

## Online supplemental material

**Fig. S1**. shows that anti-MSP1_FL_ IgG antibody levels significantly exceeded IgM levels and the magnitude of cytophilic antibodies was significantly higher compared to non-cytophilic antibodies. **Fig. S2**. shows that anti-MSP1_FL_ antibody responses are significantly correlated with antibody levels against whole merozoites. **Table S1**. Shows significantly different responses against peptides between treated and non-treated volunteers.

## Data Availability

Further information and requests for resources and reagents should be directed to and will be fulfilled by Prof. Faith Osier

## Resource availability

### Lead contact

Further information and requests for resources and reagents should be directed to and will be fulfilled by Lead Contact, Prof. Faith Osier

### Materials availability

This study did not generate new unique reagents.

### Data and code availability

The study protocol and outcomes are published (Kapulu et al., 2022; Kapulu et al., 2018). The other original data that support the findings of this study are available from the corresponding author upon reasonable request;

### Members of the CHMI-SIKA Study Team

Abdirahman I. Abdi^3^, Yonas Abebe^7^, Philip Bejon^3,8^, Peter F. Billingsley^7^, Peter C Bull^10^, Zaydah de Laurent^3^, Mainga Hamaluba^3^, Stephen L. Hoffman^7^, Eric R. James^7^, Melissa C. Kapulu^3^, Silvia Kariuki^3^, Domitila Kimani^3^, Rinter Kimathi^3^, Sam Kinyanjui^3,9,11^, Cheryl Kivisi^11^, Johnstone Makale^3^, Kevin Marsh^3,8^, Khadija Said Mohammed^3^, Moses Mosobo^3^, Janet Musembi^3^, Jennifer Musyoki^3^, Michelle Muthui^3^, Jedidah Mwacharo^3^, Kennedy Mwai^3,4^, Francis Ndungu^3^, Joyce M. Ngoi^3^, Patricia Njuguna^3^, Irene N. Nkumama^2,3^, Omar Ngoto^3^, Dennis O. Odera^3^, Bernhards Ogutu^9,12^, Fredrick Olewe^9^, Donwilliams Omuoyo^3^, John Ong’echa^9^, Faith H. A. Osier^3,6^, Edward Otieno^3^, Jimmy Shangala^3^, Betty Kim Lee Sim^7^, Thomas L. Richie^7^, James Tuju^3,5^, Juliana Wambua^3^, Thomas N Williams^3,13^.

### Affiliations

^7^Sanaria Inc., Rockville, MD, USA

^8^Centre for Tropical Medicine and Global Health, Nuffield Department of Medicine, University Oxford, Oxford, UK

^9^Centre for Clinical Research, Kenya Medical Research Institute, Kisumu, Kenya

^10^Department of Pathology, University of Cambridge, Cambridge, UK

^11^Pwani University, P. O. Box 195-80108, Kilifi, Kenya

^12^Center for Research in Therapeutic Sciences, Strathmore University, Nairobi, Kenya

^13^Department of Medicine, Imperial College London, London, UK

## Acknowledgements

We are grateful to all the study volunteers who have participated in the CHMI-SIKA study. We are also very grateful to the study teams at the study sites in Kilifi and Ahero, the collaborating teams at Sanaria, the study investigators, and all the clinical and laboratory teams. The CHMI-SIKA study was supported by a Wellcome Trust grant (107499) and sponsored by the University of Oxford. This work was supported by a Sofja Kovalevskaja Award from the Alexander von Humboldt Foundation (3.2 − 1184811 -KEN -SKP) and an EDCTP Senior Fellowship (TMA 2015 SF1001) which is part of the EDCTP2 programme supported by the European Union awarded to F.H.A.O. K.M was supported by an NIHR Global Health Research Unit grant number 16/136/33; Tackling Infections to Benefit Africa (TIBA). K.M was also supported through the DELTAS Africa Initiative Grant No.107754/Z/15/Z-DELTAS Africa SSACAB and from DELTAS Africa Initiative [DEL-15-003]. The DELTAS Africa Initiative is an independent funding scheme of the African Academy of Sciences (AAS)’s Alliance for Accelerating Excellence in Science in Africa (AESA) and supported by the New Partnership for Africa’s Development Planning and Coordinating Agency (NEPAD Agency) with funding from the Wellcome Trust [107769/Z/10/Z] and the UK government. The views expressed in this publication are those of the author(s) and not necessarily those of AAS, NEPAD Agency, Wellcome Trust or the UK government We also thank Prof. Michael Lanzer and Sumaya Biotech for providing recombinant antigens.

## Author contribution

F.H.A.O. and M.R. conceived the study and wrote the manuscript with contributions from I.N.N., S.K., M.B., K.M., D.O., J.T., K.F., R.F, E.C., M.C.K. The experiments and the analysis were performed by M.R., I.N.N., K.W. M.R. prepared the figures. All authors read and approved the final version of the manuscript.

