## Supplementary Figures for "Full-length merozoite surface protein 1 of *Plasmodium falciparum* is a major target of protective immunity following controlled human malaria infections"

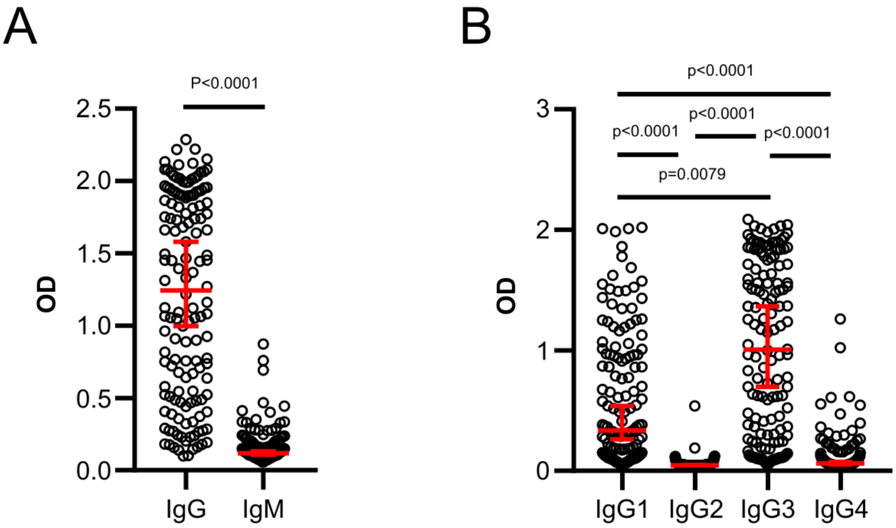


**Figure S1: The levels of IgG and cytophilic antibody responses are higher compared to IgM and non-cytophilic antibodies.**
(**A**) Comparison of IgG and IgM levels in CHMI volunteers (n=142). (**B**) Comparison of IgG subclass 1-4 antibody levels in CHMI volunteers. Each data point represents antibody levels for one sample in duplicates. Error bars represent the median plus 95% confidence intervals. Statistical differences between IgG and IgM were calculated using Mann-Whitney test and between subclass 1-4 antibodies using Kruskal Wallis test with Dunn's multiple comparisons test.

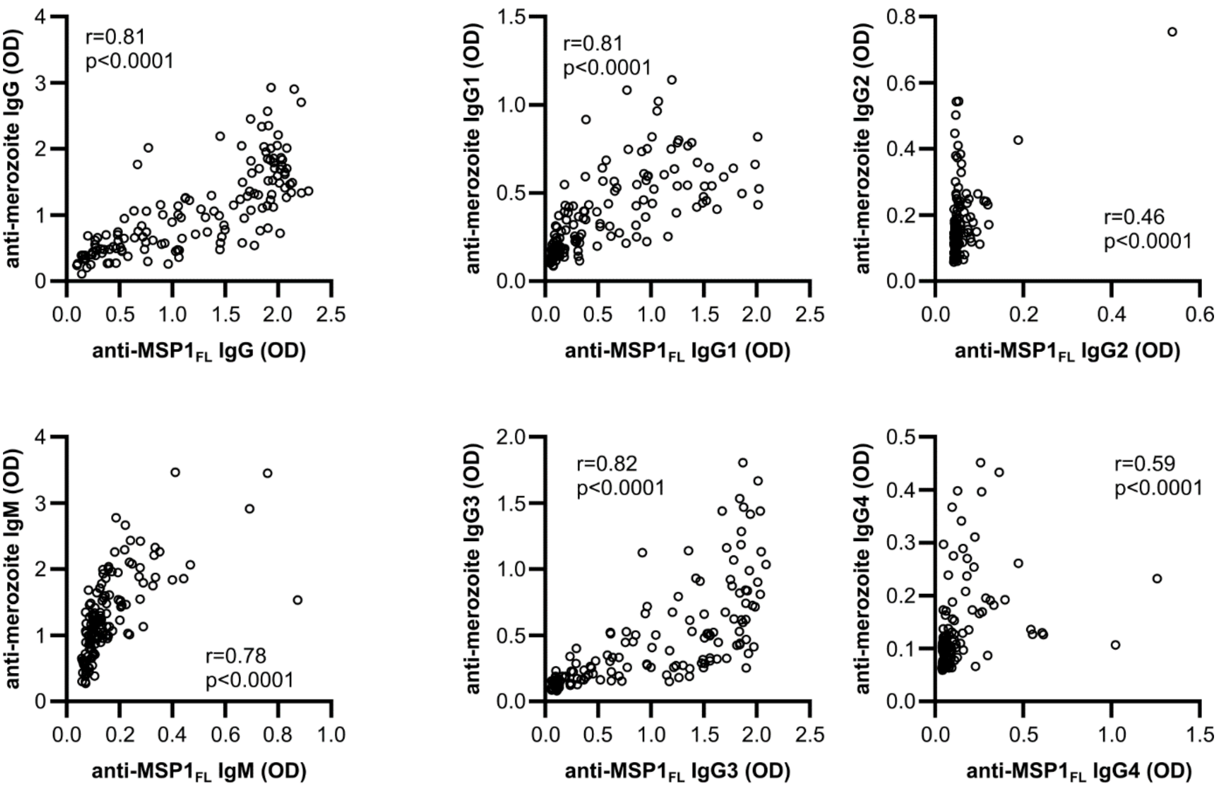


**Figure S2: Antibody responses against $MSP1_{FL}$ are correlated with responses against whole merozoites.**

Spearman's correlation of anti-$MSP1_{FL}$ and anti-merozoite IgG, IgM and IgG subclass 1-4 antibodies in CHMI volunteers (n=142). Each data point represents antibody levels for one sample in duplicates.
